# Risk stratification as a tool to rationalize quarantine among health care workers exposed to COVID-19 cases – Evidence from a tertiary healthcare centre in India

**DOI:** 10.1101/2020.07.31.20166264

**Authors:** Ravneet Kaur, Shashi Kant, Mohan Bairwa, Arvind Kumar, Shivram Dhakad, D Vignesh, Aftab Ahmad, Pooja Pandey, Arti Kapil, Rakesh Lodha, Naveet Wig

## Abstract

**Background:** Quarantine of healthcare workers (HCWs) exposed to COVID -19 confirmed cases is a well-known strategy for limiting the transmission of infection. However, there is need of evidence-based guidelines for quarantine of HCWs in COVID -19.

**Methods:** We describe our experience of contact tracing and risk stratification of 3853 HCWs who were exposed to confirmed COVID-19 cases in a tertiary health care institution in India. We developed an algorithm, on the basis of risk stratification, to rationalize quarantine among HCWs. Risk stratification was based on the duration of exposure, distance from the patient, and appropriateness of personal protection equipment (PPE) usage. Only high-risk contacts were quarantined for 14 days. They underwent testing for COVID-19 after five days of exposure, while low-risk contacts continued their work with adherence to physical distancing, hand hygiene, and appropriate use of PPE. The low-risk contacts were encouraged to monitor for symptoms and report for COVID-19 screening if fever, cough, or shortness of breath occurred. We followed up all contacts for 14 days from the last exposure and observed for symptoms of COVID-19 and test positivity.

**Results and interpretation:** Out of total 3853 contacts, 560 (14.5%) were categorized as high-risk contacts, and 40 of them were detected positive for COVID-19, with a test positivity rate of 7.1% (95% CI = 5.2 – 9.6). Overall, 118 (3.1%) of all contacts tested positive. Our strategy prevented 3215 HCWs from being quarantined and saved 45,010 person-days of health workforce until June 8, 2020, in the institution.

We conclude that exposure-based risk stratification and quarantine of HCWs is a viable strategy to prevent unnecessary quarantine, in a healthcare institution.

**Summary:** *What is already known about this subject?:*

- Quarantine of HCWs is a well-known strategy for community and HCWs to prevent the transmission of COVID-19.
- Though success stories of prompt contact tracing and quarantine to control COVID-19 are available from countries like South Korea, Singapore, and Hong Kong, there is a scarcity of evidence that could guide targeted quarantine of HCWs exposed to COVID -19 in India.

*What does this study add?:*

- Only 14.5% HCWs exposed to COVID-19 cases were stratified “high risk” contacts, and the most common reason for high-risk contacts was non-formal workplace interactions such as having meals together.
- The overall test positivity rate among the high-risk contacts was 7.1%, while it was higher in symptomatic high-risk contacts as compared to those who were asymptomatic (10.2% vs. 6.3%).

*How might this impact on clinical practice?:*

- Contact tracing and risk stratification can be used to minimize unnecessary quarantine of COVID-19 exposed health care workers and prevent the depletion of healthcare workers amidst the pandemic to continue the healthcare services optimally.

## Introduction

In India, the coronavirus disease (COVID-19) pandemic is on the rise ever since the first case was detected on January 30, 2020 (1). As of July 16, 2020, the numbers of COVID-19 cases crossed one million mark with more than 25,000 deaths (2). With the progression of pandemic, the risk of exposure of health care workers (HCWs) to COVID-19 patients was increasing. Transmission of COVID-19 infection to HCWs in health care settings weaken the health care delivery systems at such crucial times. It is further underscored by the high numbers of deaths of HCWs due to COVID-19 across the globe (4).

Case investigation, contact tracing, and quarantine remain an essential public health tool for controlling infectious disease outbreaks (5). The decision regarding whom to quarantine is of utmost importance. Irrational or unnecessary quarantine may lead to a shortage of workforce adversely impacting health care services. Whereas, not quarantining those at high risk of infection following COVID-19 case exposure may potentially lead to transmission of infection to other HCWs and patients. The World Health Organization (WHO) and the U. S. Centers for Disease Control and Prevention (CDC) issued interim recommendations for protection and quarantine of HCWs from time to time (6,7). Consequently, the Government of India also changed its quarantine policies (8,9). Though success stories of prompt contact tracing and quarantine to control COVID-19 are available from countries like South Korea, Singapore, and Hong Kong, there was a dearth of evidence that could guide targeted quarantine in India (10-12). In the initial phases of the pandemic, there was a constraint of testing facilities. Therefore, we developed an algorithm, which was based on risk stratification, to rationalize quarantine among HCWs. We report, in this paper, our experience of quarantining the HCWs based on contact tracing and risk stratification in a tertiary care centre in India.

## Methods

### Study setting

This study was conducted at a tertiary health care institution, namely the All India Institute of Medical Sciences (AIIMS), New Delhi, India. In the institution, the hospital premises and the patient care areas were divided into COVID-19 designated areas and non-COVID-19 areas for infection control measures. A vast majority of HCWs were employees of the institution who were covered under the Government of India’s leave rules. However, some of the sanitation staff and hospital attendants were procured from a manpower contractor. These employees, we understand, were paid only for the period for which they provided services.

The COVID areas included an out-patient screening facility, an in-patient facility for management of cases, and a critical care facility for COVID patients. The institution had a designated virology laboratory for testing of COVID-19, which was approved by the Indian Council of Medical Research. A team of expert virologists conducted the qRT-PCR for SARS Coronavirus 2. The test results were made available online within 24 hours of sample collection.

The Hospital Infection Control Committee (HICC) of the institution prepared the guidelines for infection control. It made recommendations for the use of personal protective equipment (PPE) in different areas of patient care. Adequate training of all HCWs in donning and doffing of PPE, social distancing, terminal cleaning, and routine disinfection was imparted. Nodal officers were appointed in each department to ensure the implementation of infection control practices.

### Study design

Case ascertainment-based cohort study

### Study period

April 11 to June 8, 2020

### Study participants

We included all HCWs and other employees of the institution who exposed with another person, (including patients, their attendants, and hospital staff) who had tested positive for COVID-19 at the institution.

### Contract tracing process and development of risk assessment tool

#### Central Contact Tracing Team (CCTT)

The CCTT was a multidisciplinary team, which consisted of experts from community medicine, infectious disease, hospital administration, internal medicine, and nursing. The team reviewed the existing literature on COVID-19 risk assessment, and prepared risk stratification criteria by synthesizing and adapting the guidelines of the WHO, Centres for Disease Control (CDC), and other evidence available in the scientific literature (7,13,14).

#### Protocol for Reporting of cases and Contact Tracing

Whenever a COVID-19 positive case was reported from any area of the institution, the concerned doctor-in charge or nodal officer at the respective department contacted the CCTT. A central helpline telephone number for CCTT was also created. The affected employee could also directly report to CCTT by calling on this helpline number. The CCTT carried out epidemiological investigation for all COVID-19 positive cases. The period for the investigation started from 48 hours before the onset of the symptoms among symptomatic cases or 48 hours before the collection of nasopharyngeal swab among asymptomatic cases. It ended at the date when COVID-19 test results became available. We interviewed the cases and checked their duty roster for preparing the list of contacts. Information from all the listed contacts was gathered either through face-to-face or telephonic interview.

#### Procedure for testing and follow up of contacts

The high-risk contacts were advised quarantine for 14 days and tested at the institution on the fifth day after exposure. All HCWs categorized as low risk but who became symptomatic during 14 days of follow up also underwent COVID-19 testing. The low-risk asymptomatic contacts continued to work, with adherence to standard precautions. All contacts were followed for 14 days after the last day of exposure to the COVID19 case.

### Operational definitions

#### COVID-19 confirmed case

A person who tested positive for COVID-19 by qualitative real-time reverse transcriptase-polymerase chain reaction (qRT-PCR) test, conducted at the virology laboratory of the institution, or at any other laboratory approved by the Indian Council of Medical Research (ICMR).

#### Risk Stratification

##### High-risk contacts

Contact with a confirmed case with any of the following:

1. Performed a respiratory Aerosol Generating Procedure (AGP) without ANY one of the following – N95 face mask, eye/face protection, gloves.
2. If the patient’s body fluids, or respiratory tract secretions, or saliva came in contact with non-intact skin or mucous membrane.
3. Present within 1 meter of the confirmed case without a mask (both case and contact) for more than 15 minutes.
4. Household contacts of a known positive case

##### Low-risk contacts

All contacts other than “high risk” contacts were classified as low-risk contacts.

##### Symptomatic contact

Any contact having symptoms suggestive of COVID-19, such as fever, cough, or breathing difficulty at the time of risk assessment.

##### Asymptomatic contact

Any contact without COVID-19 symptoms such as fever, cough, or breathing difficulty when risk was assessed, irrespective of the development of symptoms later during 14 days follow-up period.

### Statistical Analysis

Data were entered and managed in MS Excel 2016, and statistical analysis was carried out using STATA 12.0 (Stata Corp LLC 4905, Texas, USA). Results were presented as frequencies or proportions and 95% confidence interval (CI).

### Ethical Considerations

The Institute Ethics Committee approved the study. Informed verbal consent was taken on the phone from the participants. The information collected was kept confidential and unique identifiers were removed before analysis.

## Results

A total of 321 COVID-19 positive cases were reported to the CCTT during the study period. Of the 321 cases, 238 (74.1%) were HCWs and other employees, while 83 (25.9%) were patients admitted in non-COVID areas due to other medical conditions. Among the COVID-19 positive HCWs, the largest proportion was contributed by hospital attendants and sanitation workers (n = 84; 35.3%), followed by nurses (n=41; 17.2%), and security personnel (n=32; 13.3%). Doctors and laboratory staff contributed similar proportion i.e. approximately 12.2% (n= 29) each. Others included administrative staff (n = 13 (5.5%), and staff like driver, cook, food bearer etc. (n = 11; 4.6%).

The CCTT tracked 3,853 hospital employees who exposed to these 321 COVID-19 cases, both patients as well as other HCWs, during the period from April 11 to June 8, 2020 *(Figure 1)*. On risk stratification, 560 (14.5%) contacts were classified as “high risk” and 3,293 (85.5%) as “low risk” contacts. The mean number of contacts for an index case was 13.5. The maximum number of contacts associated with a single case was 87, out of which 21 (24%) were high risk.

**Figure 1.**
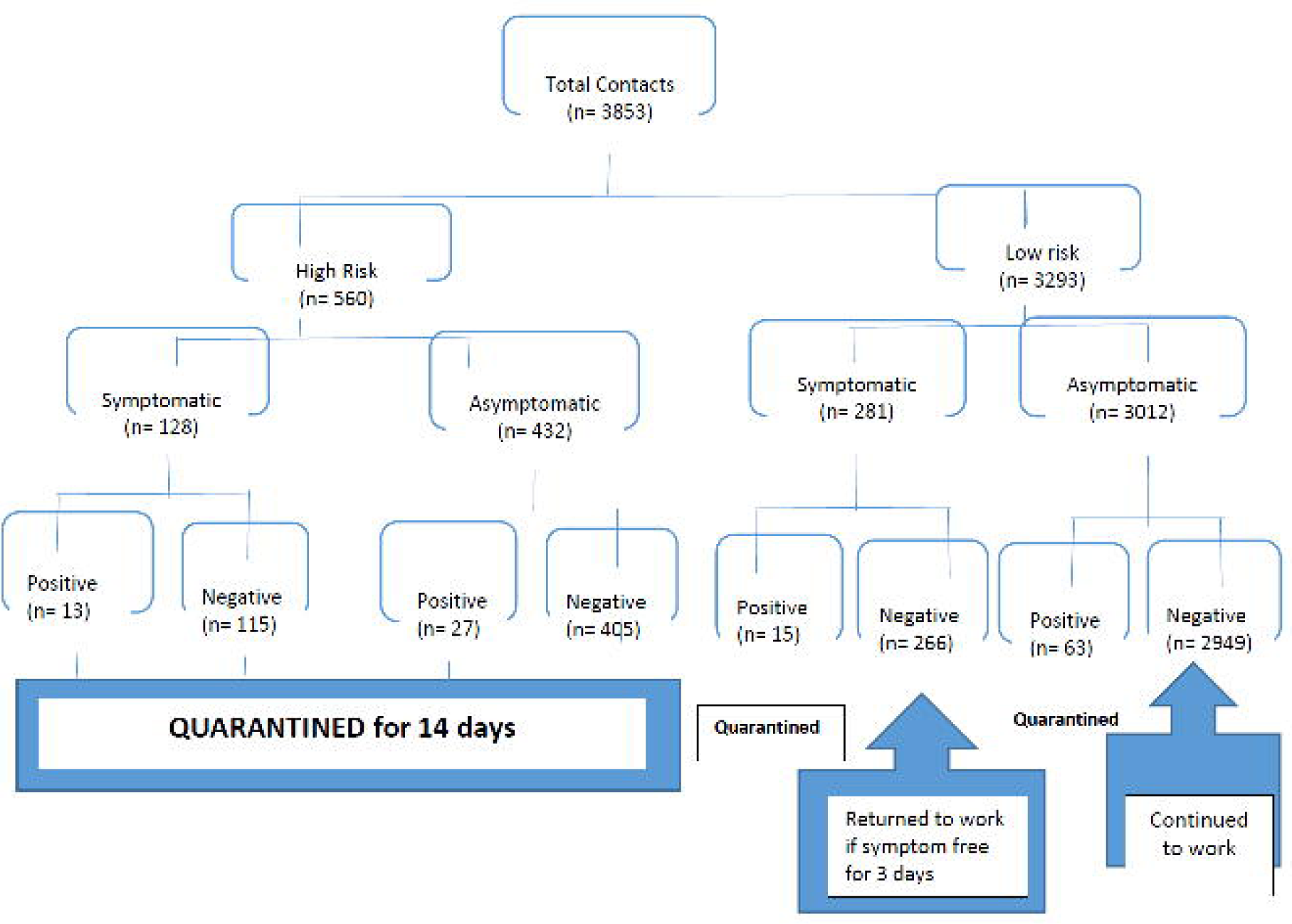
Flow diagram of contacts to COVID 19 cases follow up and quarantine.

The two most common reasons for high-risk exposure (∼40% each) were informal workplace interactions (not related to patient care), and during routine work being without mask and not maintaining physical distance. Other reasons included patient care without appropriate PPE, cleaning, or housekeeping activities without adequate PPE *(Table 1)*. In some cases, there were multiple reasons for high-risk exposure. In the patient care settings, the high-risk exposure was largely due to non-use of appropriate PPE, e.g., performing aerosol-generating procedures without eye/face protection, not performing hand hygiene after the examination or inconsistent use of N 95 mask.

**Table 1:** Distribution of reason for high-risk exposures in HCWs, AIIMS, New Delhi.

| **Reason** | **Number**<br>**(n=560)** | **Percentage** |
| --- | --- | --- |
| Informal workplace interactions<br>(Food or Tea together) | 223 | 39.8 |
| Not maintaining physical distancing and not using a<br>mask during routine work | 222 | 39.6 |
| Direct patient care without appropriate PPE | 100 | 17.9 |
| Cleaning or Sanitation activities without appropriate PPE | 14 | 2.5 |
| Household contacts of a positive patient | 11 | 2.0 |

\*Multiple reasons in some cases

Of the total 3,853 contacts, 118 (3.1%) tested positive for COVID-19 by RT-PCR. Among the 560 high-risk contacts, 40 (7.1%; 95% CI 5.2– 9.6) became COVID-19 positive during the quarantine period. The positivity rate among the symptomatic high-risk contacts was higher (10.2 %; 95% CI 5.5 – 16.7) compared to those who were asymptomatic (6.3%, 95% CI 3.9 – 8.5). Similarly, the positivity rate among the low-risk symptomatic contacts was higher (5.3%, 95% CI 3.0 – 8.7) compared to low-risk asymptomatic contact (2.1% (95% CI 1.6 − 2.6) *(Table 2)*.

**Table 2:** Test positivity rate of COVID-19 infection among the contacts, by symptoms, AIIMS, New Delhi.

| **Risk Category**<br>**(n=3853)** | **Symptom status**<br>**n (%)** |  | **Positive test result**<br>**n (%), 95%CI)** |
| --- | --- | --- | --- |
| High-risk<br>(n=560) | Symptomatic | 128 (22.9) | 13 (10.2, 5.5- 16.7) |
|  | Asymptomatic | 432 (77.1) | 27 (6.3, 4.2- 9.0) |
| Low-risk*<br>(n =3293) | Symptomatic | 281 (8.5) | 15 (5.3, 3.0- 8.7) |
|  | Asymptomatic | 3012 (91.5) | 63 (2.1, 1.6- 2.6) |
| **TOTAL** | **3853** |  | **118 (3.1, 2.5 - 3.7)** |

## Discussion

We followed up a total of 3,853 contacts, i.e., the HCWs who were exposed to COVID-19 cases. Most of the contacts (85.5%) had “low risk” exposure as per the risk stratification criteria. During the 14 days of the quarantine period, 118 contacts (3.1%, 95% CI 2.5 – 3.7) became COVID-19 positive. The test positivity rate is a proxy indicator for secondary attack rate (SAR) of COVID-19. It is higher in our study than the SAR (1.56%, 95% CI = 0.73 – 2.93) reported by nationwide contact tracing data from Taiwan (14). This indicates the relatively rapid progression of COVID-19 pandemic in our institution.

We observed a clear trend in the positivity rate with the highest rate among high-risk symptomatic contacts and lowest among low-risk asymptomatic contacts. Still, the test positivity rate among low-risk asymptomatic contacts was 2.1%. The risk stratification was based on history. It is possible that sanitation staff and hospital attendants supplied by manpower contractors feared that if they were classified as high risk, there would be mandatory quarantine and they would lose wages for that duration. Hence, some of them might have deliberately given an incorrect history of exposure. If this were to be true, then the utility of the tool would further improve in situations where paid leave is allowed for the quarantine period.

We did not physically verify the COVID-19 test status of individual HCWs. Instead, we had assumed that anyone testing positive would be notified to the CCTT. HCWs for which no positive report was communicated to CCTT were deemed to be COVID-19 negative. However, there could be some HCWs who had tested positive but were not notified to us. Therefore, the observed positivity rate would be an underestimate. We believe that the notification system at the institution was sufficiently robust. The underestimation, if any, is likely to be small enough so as not to challenge the validity of findings seriously.

While calculating the positivity rate, we had assumed that all contacts were susceptible at the time of exposure. However, some of them might already have had protective antibodies. It would be particularly true because a substantial proportion of COVID infection is reported to be asymptomatic. Such a situation would result in an underestimation of the positivity rate. We had assumed that none of the contacts were in the incubation period of COVID infection at the time of their exposure. If the HCW was truly in the incubation period, then s/he would have tested positive irrespective of current exposure. Under such a scenario, the observed the positivity rate would be an overestimate of the true situation. We could not find any published literature that could assist in estimating the quantum of such overestimate. However, we feel that the number of HCWs that could have been in the incubation period of COVID infection is likely to be small and not vitiate the findings of this study.

Through the risk stratification tool, we aimed to identify those exposed HCWs that required quarantine to protect patients and other HCWs from potential infection. Simultaneously, we also aimed to avoid unnecessary quarantine of those HCWs that were likely to be uninfected, and thereby prevent workforce depletion. Incorrectly labelling an HCW as low risk and thereby allowing her/him to continue to work rather than being quarantined, entailed serious consequences to co-workers and patients. It was preferable to err on the side of caution at the cost of loss of workforce. The test positivity rate among low risk symptomatic (5.3%) was closer to the high-risk group (7.1%) compared to low risk asymptomatic (2.1%). If we convert the test positivity rate among low-risk asymptomatic contacts in numbers, it will be a substantial number. These asymptomatic contacts may further transmit the infection to others, which is evident from the literature (15). The asymptomatic transmission of COVID-19 remains a major challenge for the protection of HCWs and the curb of pandemic progression. Thus, all low-risk asymptomatic contacts were advised to observe adherence to precautions such as physical distancing, hand hygiene and PPE as appropriate, so that in case of an error in stratification, they do not pose the risk of transmission to others and avoid further resurgence of COVID-19 cases.

We found that informal interactions such as having snacks/meals together was one of the most common reasons for high-risk exposure among the contacts. Our findings support the WHO guidelines that had emphasized the exposure in the common dining facilities (16). We, therefore, suggest that an appropriate dining facility that would enable maintenance of proper physical distance should be provided at health care facilities. Other measures to avoid overcrowding may include restricting the number of persons inside the dining room at any given point of time. Staggering the lunch timings could be yet another such measure. We also recommend that during training program of the HCW for infection control practices, the risk due to dining together may be emphasized.

Our risk stratification strategy prevented the depletion of human resources for continuing healthcare services. In our study, 560 contacts were high risk. They were eligible quarantine for 14 days. Of the low-risk contacts (3293), 78 tested positive and thus quarantined. Therefore, this strategy prevented 3215 HCWs from quarantine following exposure and saved 45,010 person-days till June 8, 2020. In contrast, all contacts irrespective of the risk status were quarantined in most healthcare institutions, particularly in the facilities with constraint resources for testing. It led to severe depletion of the workforce, and subsequently, shut down of many healthcare institutions in worse affected cities, Mumbai and Delhi (17–20).

### Strengths

We employed multiple strategies, including personal interview and review of records, to prepare the list of contacts of exposed HCW. We, therefore, feel that all potential contacts were listed and tracked. Thus, the data quality was expected to be satisfactory. The testing for COVID infection was done at an accredited laboratory. All quarantined participants were contacted, and history obtained at the end of 14 days of follow up.

### Limitations

The risk stratification was based on the history provided by the participant. Recall and social desirability bias, therefore, could not be ruled out. Deliberate misinformation was also a possibility, particularly where participants feared that if quarantined, they could lose wages for that period. Sanitation staff and hospital attendants that were provided by an external agency were more likely to fall into this category.

## Conclusions

The risk stratification tool developed by us served its intended purpose of correctly categorizing the risk category of exposure, as well as minimizing unnecessary quarantine of COVID-19 exposed health care workers. The utility of the tool could be further enhanced by some modification in existing quarantine policy.

## Data Availability

Data are available on request. Deidentified data will be made available on request from a bona fide research institute or entity for use in a valid research study. Requests should be made via email to Ravneet Kaur.

## Declarations

### Contributors

RK, SK, and MB conceptualized the paper. RK, MB, and AK coordinated the contract tracing and risk stratification. MB and RK developed the analysis plan. MB, RK, SK and VD implemented the data analysis, and RK, SK, AK, and RL contributed to data interpretation. RK drafted the majority of the manuscript, and MB drafted sections of the manuscript. AK, RL, AS, NW, SD, PP, and AA critically reviewed and provided comments on the first draft of the manuscript. All authors read and approved the paper.

### Patient and public involvement

Consent has been taken from all the study participants.

### Patient consent for publication

Not required.

### Ethics approval

The ethical clearance for this study was obtained by the Institutional Ethics Committee of AIIMS, New Delhi.

### Provenance and peer review

Not commissioned; externally peer-reviewed.

## Acknowledgments

We acknowledge the help of department nodal officers of the Hospital Infection Control Committee and resident doctors of AIIMS, New Delhi, in contact tracing of COVID-19 positive cases.

